# Reinventing the Body Mass Index: A Machine Learning Approach

**DOI:** 10.1101/2024.04.26.24306457

**Authors:** JM Peregrin-Alvarez

**Affiliations:** Freelance PhD Scientist and CEO at Virtual Personal Trainer, Spain

**Keywords:** Body Mass Index, BMI, BMI Classification, Obesity, Weight Management, Health, Machine Learning, ML, Prediction

## Abstract

This study explores the predictive capabilities of the Body Mass Index (BMI) formula across a diverse dataset, examining the potential enhancements achievable through integrating additional parameters using machine learning (ML) models. Various modern ML models were utilized (K-Nearest Neighbors, Neural Networks, Decision Trees, Support Vector Classification, Logistic Regression, and Ridge Classifiers. Ensemble models: voting Classifier, Random Forest, and Gradient Boosting), demonstrating improved accuracy and precision over the traditional BMI calculations. Incorporating age and gender into BMI calculations together with the best performing ML model such as Gradient Boosting offers promise for more accurate and personalized health assessments, with significant implications for clinical practice and public health interventions.

## Introduction

The Body Mass Index (BMI) has long been a staple metric for assessing body weight relative to height in health assessments [1]. However, some studies have highlighted its limitations, prompting exploration into alternative approaches. For example, due to the variety of body types, muscle distribution, bone mass, etc, BMI is not appropriate as the only indication for diagnosis, which could lead to misclassification [2]. However, weight control is a key factor in the prevention of non-communicable diseases. Recent studies have shown the utility of Machine Learning (ML) in clinical settings. For example, a recent ML approach predicted weight changes over the years, which could be helpful for weight management approaches [3]. Thus, it will be interesting to evaluate the efficacy of the traditional BMI formula and investigate the potential improvements offered by modern ML classification models by incorporating additional parameters other than the traditional height and weight. For example, as a person ages, body fat mass naturally increases, and muscle mass declines. Numerous studies have shown that a higher BMI of 23.0–29.9 in older adults can be protective against early death and disease [4]. Other studies have indicated that the risk for heart disease and diabetes increases in women with a waist measurement greater than 35 inches (88.9 cm) and more than 40 inches (101.6 cm) in the case of men [5]. Furthermore, the BMI may not accurately reflect the health of certain racial and ethnic populations. For example, numerous studies have shown that people of Asian-Pacific descent have an increased risk of chronic disease at lower BMI cut-off points, which leads to specific BMI guidelines with alternative BMI cut-off points for this population [6].

This paper investigates the potential of modern ML classification models to enhance BMI calculations by incorporating additional parameters such as age, gender, and ethnicity. Leveraging data from the National Health and Nutrition Examination Survey (NHANES) [7], we aimed to provide novel insights into BMI calculations and their implications for health assessments. NHANES is a program of studies designed to assess the health and nutritional status of adults and children in the United States, a subprogram of the Centers for Disease Control and Prevention (CDC). Survey data is intended to be used in epidemiological studies and health sciences research, which help develop sound public health policy, direct and design health programs and services, and expand the health knowledge for the Nation. These data were fundamental to conducting our comprehensive analysis aiming to provide potential new alternative measurements to the traditional BMI calculations. We also employed a wide variety of modern ML models: K-Nearest Neighbors, Neural Networks, Decision Trees, Support Vector Classification, Logistic Regression, and Ridge Classifiers. Ensemble models: voting Classifier, Random Forest, and Gradient Boosting, aiming to provide the most reliable and accurate results.

## Methods

We downloaded data from 5663 individuals obtained from surveys combining interviews and physical examinations from NHANES (years 1999 to 2022). The dataset comprises weight, height, age, gender, ethnicity, and BMI variables. Data Privacy was a priority for the NHANES dataset. Using this robust NHANES dataset, we employed various ML models (K-Nearest Neighbors [8], Neural Networks [9], Decision Trees [10], Support Vector Classification (SVC)[11], Logistic Regression [12], and Ridge Classifiers [13]. ML ensemble models: voting Classifier [14], Random Forest [15], and Gradient Boosting [16]) to predict BMI categories based on the inclusion of additional parameters such as age, gender, and ethnicity. Classification models are well-suited for predicting and assigning observations into discrete categories. Subsequently, ML classification models were employed to discern whether incorporating additional parameters such as age, gender, and ethnicity could enhance predictive accuracy. We compute the standard BMI formula and compare different ML predictions (see below) against real BMI classification values (Underweight, BMI below 18.5; Normal, BMI 18.5 – 24.9; Overweight, BMI 25.0 – 29.9, and Obese BMI 30.0 and above). Categorical variables (gender and ethnicity) were transformed into numerical representations, and all training parameters were subjected to feature importance, distribution, balance and correlation analysis to analyze the quality of the different datasets.

To evaluate model performance and measure the agreement between the predicted and actual classifications, the following statistical analyses were conducted: Accuracy, Balanced Accuracy, Precision, Recall, Precision-Recall Curve, Area Under the Curve (AUC), F1- and F-beta Scores, Matthews Correlation Coefficient (MCC), and, Cohen’s Kappa.

For ML model/s selection, the models with the highest consistent values across all statistical metrics (Average) were selected. Additionally, cross-validation techniques/scores (CVS) were employed to ensure the reliability and generalizability of our findings. Specifically, we utilized Stratified K-Fold cross-validation, which is particularly suitable for maintaining the distribution of target classes across folds. This technique involves splitting the dataset into K folds while preserving the proportion of each class in every fold. In our study, we set K to 5, resulting in five folds for training and evaluation. This approach allows us to effectively assess the generalization performance of our models and mitigate the risk of overfitting. Finally, we assessed the degree of agreement/ disagreement between the selected predictive ML model and the traditional BMI formula by employing a comprehensive analysis using 1000 random prompt instances.

## Results

Our findings indicate that the features of the traditional BMI formula, based solely on weight and height, exhibits reasonable predictive power across most ML models (Fig. 1, Tables 1 and 2). However, some ML models trained on datasets enriched with additional age and gender information were able to outperform the reference model. Most models demonstrated a high precision and accuracy, while models such as SVC, Gradient Boosting, and Neural Network, demonstrated to outperform the reference model. The results show that training the ML models with additional age and gender parameters, particularly the HWA and HWAG datasets, can notably improve the current BMI formula predictive capability.

**Table 1.**
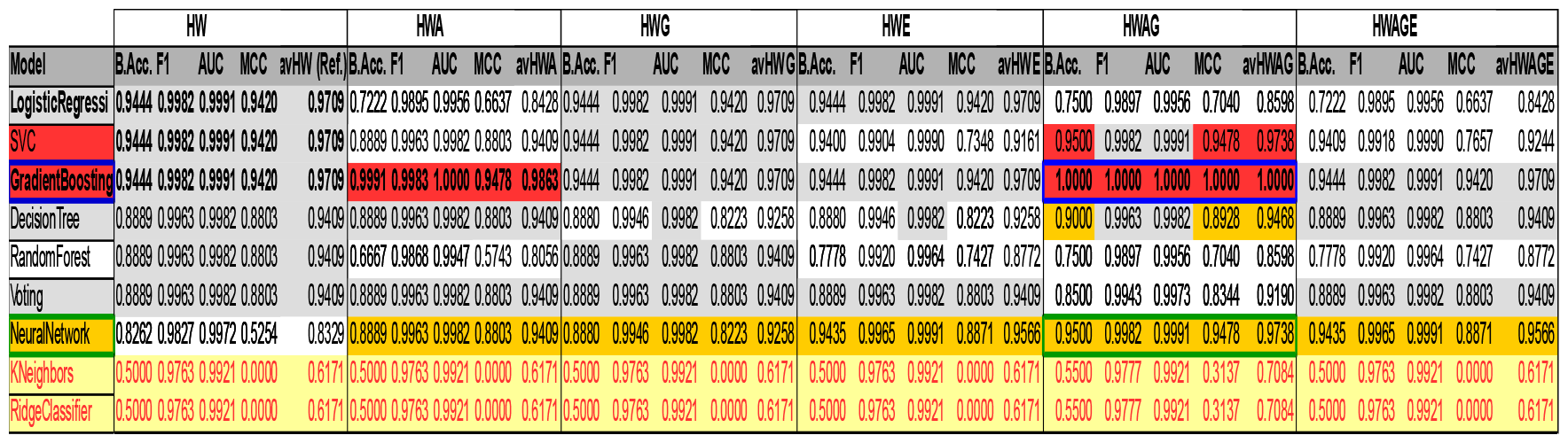
Balanced Accuracy, F1-score, AUC, MCC, and average statistics of the ML models. This table represents a more detailed statistical table for Fig. 1. The above statistics were chosen for representation purposes as they offer a good balance between capturing the overall performance and addressing the challenges posed by class imbalance (see Methods). Abbreviations: HW: height and weight; HWA: height, weight, and age; HWG: height, weight, and gender; HWE: height, weight, and ethnicity; HWAG: height, weight, age, and gender; and HWAGE: height, weight, age, gender, and ethnicity. B.Acc.: Balanced Accuracy; F1: F1-score; AUC: Area Under the Curve; and MCC: Matthews Correlation Coefficient. Light gray cells with bold characters stand for the best performance metrics/models in the reference dataset (HW); Red background cells indicate outperforming metrics, i.e. improved prediction values compared to the reference HW-model. Orange background cells indicate the best row metrics, other than red cells. Yellow background cells indicate the lowest values, i.e. models with lowest stat metrics. Red cells with bold characters and blue square borders indicate the best performing ML model compared to the reference and the top cross-validation models (see Table 2). Orange background cells with green square borders indicate the best cross-validation performing model.

**Table 2.** Cross-Validation Analysis. This table represents the CVS of the different ML models applied to the best performing dataset (HWAG)(see Fig.1 and Table 1). 1-5 columns stand for the five folds cross-validation sets for training and evaluation; avCVS stands for the average of the 5 folds columns; Voting3 stands for the voting ensemble model trained with the 3 best performing ML models according to Fig. 1 and Table1 (SVC, GradientBoosting, and NeuralNetwork); and Voting2 stands for the voting ensemble model trained with the 2 best performing ML models (GradientBoosting and NeuralNetwork).

| CVS(HWAG) | 1 | 2 | 3 | 4 | 5 | avCVS |
| --- | --- | --- | --- | --- | --- | --- |
| NeuralNetwork | 0.9912 | 0.9561 | 0.9735 | 0.9735 | 0.9558 | 0.9700 |
| GradientBoosting | 0.9386 | 0.9474 | 0.8850 | 0.9381 | 0.8761 | 0.9170 |
| DecisionTree | 0.9123 | 0.9298 | 0.8673 | 0.9381 | 0.8673 | 0.9029 |
| Voting | 0.9474 | 0.9474 | 0.9115 | 0.8673 | 0.9115 | 0.9170 |
| Voting3 | 0.9386 | 0.9298 | 0.8938 | 0.8938 | 0.9292 | 0.9170 |
| Voting2 | 0.9474 | 0.9298 | 0.8850 | 0.8673 | 0.9027 | 0.9064 |
| SVC | 0.8421 | 0.8333 | 0.8407 | 0.8761 | 0.8407 | 0.8466 |
| RandomForest | 0.9386 | 0.9386 | 0.9027 | 0.9027 | 0.9469 | 0.9259 |
| LogisticRegression | 0.9386 | 0.8860 | 0.8761 | 0.8230 | 0.8496 | 0.8746 |
| KNeighbors | 0.9386 | 0.9649 | 0.8938 | 0.8938 | 0.8850 | 0.9152 |
| RidgeClassifier | 0.9386 | 0.8947 | 0.8850 | 0.9204 | 0.8761 | 0.9029 |

**Figure 1.**
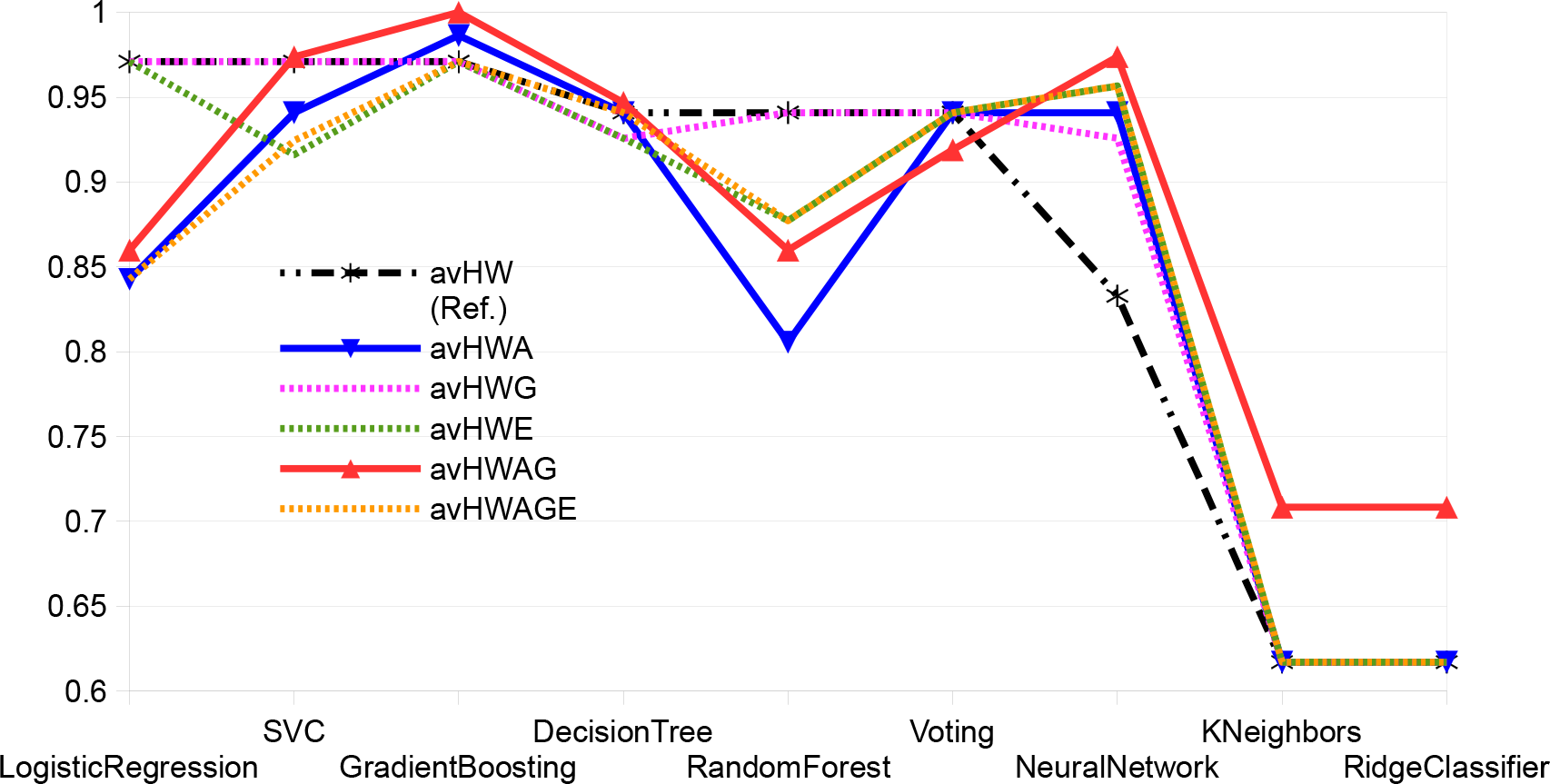
ML models performance. This graphic represents the average of all statistical measurements (see methods) from all ML models under study. Abbreviations: HW: height and weight; HWG: height, weight, and gender; HWA: height, weight, and age; HWE: height, weight, and ethnicity; HWAG: height, weight, age, and gender; and HWAGE: height, weight, age, gender, and ethnicity.

The cross-validation analysis further supports the use of NeuralNetwork and Gradient Boosting models as best performing ML models trained with additional age and gender features (Table 2).

Finally, the model performance analysis revealed a notable agreement between the Gradient Boosting model and the traditional BMI formula (Figure 2). The analysis exhibited high concordance rates, with a total of 871 agreements (75 instances demonstrating agreement between the BMI formula and the Gradient Boosting model trained only with height and weight features (HWagr), 13 instances demonstrating agreement between the BMI formula and the Gradient Boosting model trained with additional age and gender features (AGagr), and 783 instances demonstrating agreement between the BMI formula and both ML dataset-models (ALLagr). A total of 129 novel predictions were also obtained, representing the disagreement between the BMI formula and the ML models but the agreement between both ML models. Interestingly, zero disagreements were found, i.e. no disagreement between both ML models and no disagreement of any of the models with the BMI formula.

**Figure 2.**
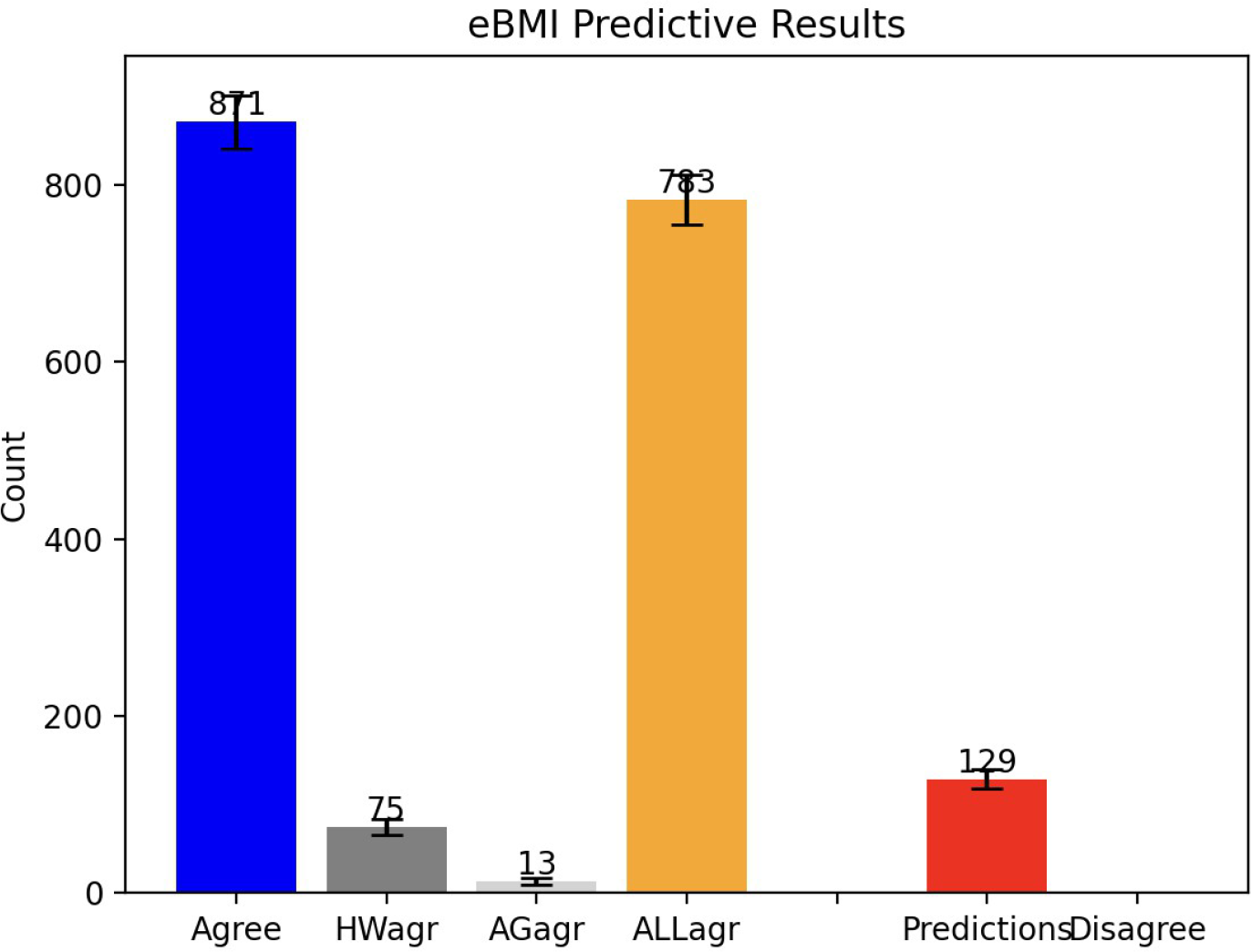
Model performance analysis. This graphic represents the performance of the Gradient Boosting ML model vs the traditional formula. Abbreviations: Agree: Total ML models agreements vs the BMI formula. HWagr: height and weight model agreement with BMI formula; AGagr: height, weight, age and gender model agreement with BMI formula; ALLagr: Both ML Models agreement and BMI formula agreement; Predictions: BMI formula disagreement but ML models agreement; Disagree: ML models disagreement and BMI formula disagreement.

In summary, our findings support the use of modern ML methods such as Gradient Boosting to: 1) support the traditional BMI results; and 2) enhance its predictive power by formulating new hypothesis. Incorporating age and gender into the traditional BMI calculations can enhance the predictive power of the formula, evidenced by the capability of the ML model to outperform the results of the traditional BMI formula. Notably, models trained with additional parameters (age and gender, apart from height and weight) demonstrated improved precision and accuracy, offering a more nuanced approach to BMI measurement. Cross-validation analysis further supported the efficacy of certain ML models such as Gradient Boosting for enhancing the predictive capability of the traditional BMI formulation.

## Discussion

BMI is a standard health assessment tool in most healthcare facilities. Although, for decades, the BMI has been widely used as a standard measurement for health based on body size, it has been criticized for its oversimplification of the real meaning of being healthy. Many researchers have claimed that BMI is outdated and inaccurate, and, perhaps, it should not be used in medical and fitness settings. For example, in epidemiological studies, the BMI based on self-reported height and weight (self-reported BMI) is subjected to measurement error [17]. Other studies have suggested adjusting the Normal BMI values to avoid false positive/ negative assignments [18]. It is expected that medical professionals would take the BMI result and consider patients as unique individuals.

However, some health professionals use only BMI to measure a person’s health status before providing medical recommendations. This can lead to weight bias and poor quality healthcare [19, 20]. Moreover, serious medical issues might go unnoticed or incorrectly seen as weight-related problems [19]. Other studies have shown that the higher a person’s BMI is, the less likely the person will attend regular health checkups due to fear of being judged, distrust of the healthcare professional, or a previous negative experience. This can lead to late diagnoses, treatment, and care [21]. However, because of the ease and efficiency of gathering height and weight information, it remains important to assess the extent of error present in self-reported BMI measures and to explore possible adjustment factors. Our study provides such possible adjustments by incorporating additional features such as age and gender and enhancing the power of BMI calculations by means of the last state-of-the art ML technologies.

The implications of our findings for clinical practice and public health interventions are substantial. By incorporating age and gender into BMI calculations, we can offer more personalized health assessments, leading to improved preventive strategies and interventions. For instance, considering age-related changes in body composition and metabolism, as well as gender-specific health disparities, could lead to more accurate assessments of obesity-related risks and improved preventive strategies. Additionally, by acknowledging the diversity in body compositions across different demographic groups, interventions can be better tailored to address the unique needs of individuals, ultimately enhancing the effectiveness of weight management programs and health interventions.

However, limitations such as dataset representativeness and practical challenges in implementing ML-enhanced BMI calculations must be addressed. While our analysis leveraged a comprehensive dataset from the National Health and Nutrition Examination Survey (NHANES), the representativeness of this dataset may be subject to certain biases, particularly in terms of ethnic diversity. As NHANES primarily represents the U.S. population, the generalizability of our findings to more diverse populations globally may be limited. For example, the specific BMI cut-off points for the Asian-Pacific population [6] highlight the importance of properly stratifying ethnic data. This could explain why using ethnicity as an additional parameter for BMI class prediction did not show any significant improvement compared to the use of age and gender. Future studies should aim to address these limitations by incorporating more diverse and representative datasets, ensuring that findings can be applied to a broader range of populations.

Furthermore, the application of some ML models such as Ridge Classifier and KNeighbors suggests some potential limitations of these models or the nature of the data used for training. Moreover, while our study demonstrates the potential of ML-enhanced BMI calculations in improving health assessments, challenges may arise in implementing these approaches in real-world healthcare settings. Practical considerations such as data accessibility, model interpretability, and integration into existing healthcare systems need to be carefully addressed. Additionally, ensuring data privacy and maintaining confidentiality in accordance with ethical guidelines should be considered. Collaborative efforts are needed to overcome these challenges and facilitate the integration of ML models into routine clinical practice.

## Conclusions

In conclusion, our study highlights the potential of ML models to enhance BMI calculations by incorporating additional parameters such as age and gender. ML models such as Gradient Boosting emerged as promising alternatives, showcasing the potential for a more nuanced and accurate approach to BMI measurement. These findings offer valuable insights into the future of BMI assessment and underscore the need for further research to refine and validate these approaches. In summary, this study provides valuable insights into the predictive capabilities of ML models for BMI classification. Our results underscore the potential for improved BMI measurements by adapting traditional formulas with additional parameters, such as age and gender. Indeed, some studies highlighted the significance of incorporating age and gender into BMI calculations for more accurate assessments of obesity and related health risks [22,23].

Future research in this domain could contribute to developing more personalized and accurate health assessment tools. The BMI only considers weight and height as a measure of health status, rather than the person. Our results suggest that considering height, weight, age and gender, and potentially other factors that may affect an individual weight and health status, such as a more comprehensive ethnicity dataset, could complement traditional BMI calculations and provide more consistent health statements. For example, health practitioners could train and test their historical patient data with the above ML models and additional BMI parameters (age, gender, or both independently), and, either serve as a validation of the results, if there is no difference with the traditional BMI results, or, in case of discrepancy, study in more detail the potential causes behind the differences between the traditional BMI formula versus the ML model prediction. Furthermore, additional data, such as body composition [24], medical history [25], and demographic and socioeconomic information [26], could help health practitioners, researchers, and scientists to provide more realistic weight management and health assessments, as well as early diagnoses, treatments, better healthcare, as well as new opportunities for R&D and scientific discovery.

In summary, our study contributes to the ongoing discourse surrounding BMI calculations and their implications for health assessments. By integrating age and gender into BMI calculations using ML techniques, we offer a novel approach to improving health assessments and tailoring interventions. Our findings underscore the potential of ML-enhanced BMI calculations in providing more personalized and accurate health assessments, ultimately leading to better health outcomes for individuals. Future research directions include exploring the influence of ethnicity on BMI calculations, refining ML algorithms for enhanced prediction accuracy, integrating additional health parameters into BMI assessments, conducting validation studies across diverse populations, and addressing ethical and practical considerations for the implementation of ML-enhanced BMI calculations in healthcare settings. Additionally, exploring the impact of other factors such as body composition, medical history, and socioeconomic status on BMI calculations could provide valuable insights into developing more comprehensive health assessment tools. By leveraging modern ML techniques, we can revolutionize BMI calculations and improve health outcomes on a global scale.

## Data Availability

All data produced in the present study are available upon reasonable request to the authors

